# Association between motor cortex grey matter loss and inability to control an ECoG-based implanted Brain-Computer Interface in ALS

**DOI:** 10.64898/2026.06.23.26355654

**Authors:** Mathijs Raemaekers, Simon H. Geukes, Erik J. Aarnoutse, Mariana P. Branco, Zac V. Freudenburg, Anouck Schippers, Nathan E. Crone, Sasha Leinders, Julia Berezutskaya, Nick F. Ramsey, Mariska J. Vansteensel

## Abstract

**Background:** The field of implantable Brain-Computer Interfaces (iBCIs) is rapidly advancing, with individuals with amyotrophic lateral sclerosis (ALS) as key beneficiaries. However, ALS-related cortical degeneration may impair iBCI effectiveness. This study investigated whether structural magnetic resonance imaging (MRI) and functional MRI (fMRI) metrics are associated with the quality of electrocorticography (ECoG) signals critical for iBCI use.

**Methods:** Six late-stage ALS participants and 76 controls underwent T1-weighted structural MRI and task-based fMRI during right-hand movement or attempts thereof. ECoG data of ALS participants was benchmarked using ECoG data acquired in epilepsy patients. Grey matter thickness in the sensorimotor cortex and fMRI activation in the motor-hand area were measured.

**Results:** Four ALS participants showed >0.4 mm thinning in the precentral gyrus, while the postcentral gyrus was spared. ECoG signal quality was significantly associated with precentral grey matter thickness, but not with fMRI activity.

**Conclusions:** These findings suggest that presurgical assessment of precentral grey matter thickness could potentially prove useful for iBCI candidate selection in advanced ALS.

**Plain Language Summary:** People with amyotrophic lateral sclerosis (ALS) can lose the ability to move and speak, but their thinking often remains intact. Implantable brain-computer interfaces (iBCIs) can help by translating brain signals into commands for communication devices. However, ALS damages the motor cortex, which may reduce the quality of these signals. In this study, we examined brain scans and electrical recordings from six people with advanced ALS. We found that thinning of the motor cortex was linked to weaker brain signals needed for iBCI control, while functional MRI activity was less predictive. This suggests that measuring motor cortex thickness before surgery could help identify who will benefit most from an iBCI, improving treatment decisions and future clinical trials.

**Two sentence summary:** We examine presurgical MRI/fMRI and ECoG recordings from people with advanced ALS receiving implanted brain-computer interfaces. Motor cortex thinning is associated with poorer ECoG signal quality, suggesting cortical thickness may help identify candidates likely to benefit.

## Introduction

The field of implantable Brain Computer Interfaces (iBCIs) is rapidly expanding. Recent years have seen accelerated progress in the development and validation of iBCIs, with multiple academic centers and device manufacturers conducting clinical trials to evaluate these technologies in people with neurological disorders. In line with this development, the American Food and Drug Agency (FDA) has expressed the need to develop standardized outcome measures to evaluate the clinical impact of BCIs for one of the key target populations: people with Amyotrophic Lateral Sclerosis (ALS).

ALS is a progressive disorder characterized by the degeneration of upper and lower motor neurons^1^. This condition results in muscle weakness, atrophy, and ultimately paralysis, severely impacting individuals’ abilities to move their limbs and to perform functions such as speaking, swallowing, and breathing ^2^. In the advanced stages of the disease, people with ALS may enter a locked-in state, where the ability to move or communicate verbally is lost due to complete paralysis, but cognitive function remains (largely) intact ^3,4^.

Implantable BCIs can bypass the damaged motor pathways of people with ALS by directly translating electrophysiological neural signals into commands for external devices, enabling them to communicate or control assistive technologies through brain activity alone ^5^. The primary signal source of iBCI systems so far has been the sensorimotor cortex ^6^ and several studies have reported successful demonstrations of iBCI control by individuals with ALS using signals from this brain region ^7,8^. The sensorimotor cortex is ideal for decoding of neural signals due to its well-defined topographic nature ^9^, and whose activity intuitively matches to interactions with the outside world ^10^.

At the same time, ALS is known to affect the electrophysiological neural signals in the sensorimotor cortex ^11,12^. If these sensorimotor signals become low or unreliable, this will impact detection and discrimination of the brain signals by the iBCI. This will in turn impact the accuracy with which brain states are decoded, resulting in a BCI that is not consistently producing the output desired by the user. Indeed, advanced ALS has been associated with diminished iBCI performance ^13^. Given that BCI implantation and training are associated with surgical risk and user and caregiver burden, it is vital to reliably identify those iBCI implant candidates who are most likely to benefit from the functionalities an iBCI can offer, and to precisely inform these individuals on the expected iBCI performance. Moreover, the inclusion of unsuitable candidates in clinical trials might interfere with accurate assessment of the functionality of an implanted device.

Here, we assessed the value of structural and functional imaging data for the iBCI eligibility assessment of people with ALS. We analyzed imaging data collected prior to BCI implantation from people with ALS who participated in the Utrecht Neural Prosthesis (UNP) or in the Cortical Communication (CortiCom) project. We examined cortical thickness, fMRI activity, and electrocorticography (ECoG) signals in the sensorimotor cortex during (attempted) hand movements. The main focus was on the 65–95 Hz range of the high-frequency band of ECoG signals. Power changes in this range and at higher frequencies reflect the same underlying asynchronous neuronal firing, rather than distinct oscillatory processes, and have been shown to be particularly informative for iBCI control ^14^. All measures were benchmarked to data of control groups. Results from this study can help to optimize clinical trial design by enabling data-driven patient stratification, thereby facilitating the translation of iBCI technologies from research to clinical practice.

We show that four of six individuals with advanced ALS have marked thinning of the precentral gyrus relative to age-matched controls, while the postcentral gyrus is relatively spared. In implanted participants, reduced precentral grey matter thickness is associated with poorer high-frequency-band ECoG signal quality during attempted movement, whereas fMRI activity shows a less consistent relationship with ECoG signal quality. These findings indicate that presurgical assessment of precentral cortical thickness may provide a useful marker for identifying individuals with ALS who are most likely to benefit from motor-based iBCI implantation.

## Materials and Methods

### Participants

A total of 6 individuals with ALS (3 females, Table 1 in Supplementary Data 1) were enrolled in either the CortiCom (n=3; clinicaltrial.gov: NCT03567213 & NCT06207591) or UNP (n=3; clinicaltrial.gov: NCT02224469) study. These studies received ethical approval from the Institutional Review Board (IRB) of the UMC Utrecht, the Netherlands (CortiCom & UNP) or the Johns Hopkins University IRB, MD, USA (CortiCom). Participants were recruited at the UMC Utrecht (n=5) and at the Johns Hopkins Hospital (n=1). All participants except CC001 were nearing the locked-in state (see Amyotrophic Lateral Sclerosis Functional Rating Scale (ALSFRS) scores in Table 1 in Supplementary Data 1). CC003 was excluded from implantation due to findings from structural and functional imaging that indicated that electrode placement directly over areas with clear voluntary modulation of activity was not feasible. Complete absence of task-related fMRI activity led to excluding UNP006 from implantation. For these 2 participants only structural and functional MRI results are available.

**Table 1:**
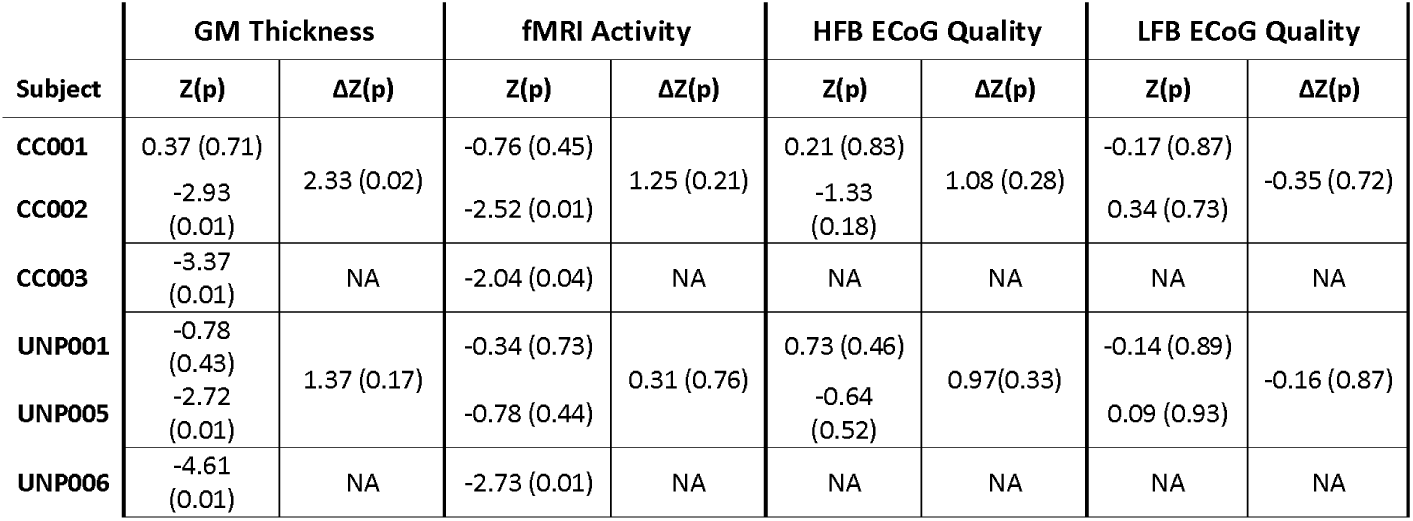
Z-values and ΔZ-values for grey matter thickness, fMRI activity and ECoG quality obtained by benchmarking to the data of the control populations. ΔZ values are calculated for the two participants using the same iBCI system (CC001 vs. CC002 and UNP001 vs UNP005). The statistical significance (two-sided, uncorrected for multiple comparisons) for each individual measurement is in brackets after the Z/ ΔZ-value.

For reference of functional and structural MRI results, we used data from a cohort of 76 healthy individuals (39 females; age range 18-89; mean/SD age; 45.7/21.7). This data was obtained as part of the creation of a diagnostic database by BrainCarta Inc (Utrecht, the Netherlands), and the acquisition was approved by the IRB of the UMC Utrecht, the Netherlands. All controls had a blank neurological history and were right-handed according to the Edinburgh handedness inventory ^15^. For reference of electrophysiological results, we used ECoG data from 24 individuals with epilepsy (12 females; age range 17-59; mean/SD age; 29.0/10.6) who were admitted to the intensive epilepsy monitoring unit of the UMC Utrecht for epileptic source localization. Of this group 11 had a left hemisphere, and 13 a right hemisphere implant.

All mentioned studies adhered to the principles outlined in the declaration of Helsinki (2013). All participants gave informed consent for participation, when necessary using a procedure tailored to people with severe motor-related communication impairments ^16^.

### Acquisition of imaging data

Functional and structural images were acquired on either a Siemens Magnetom Prisma (Siemens, Erlangen, Germany), or a Philips Achieva (Philips Medical Systems, Best, the Netherlands) 3T MRI scanner. For the ALS participants, imaging was conducted prior to BCI implantation surgery to 1) establish the participant’s ability to elicit activity in the sensorimotor cortex, 2) identify the optimal site of the implant and 3) assess the presence of any structural anomalies that could interfere with accomplishing adequate iBCI control ^17^. All ALS participants except CC001 were scanned while receiving tracheostomy invasive mechanical ventilation ^17^. The details of the MRI protocols used for ALS and control participants can be found in Table 2 in Supplementary Data 1. All protocols covered the whole brain. The fMRI task required participants to either execute (control participants and CC001 who had residual hand control) or attempt right-hand movements (ALS participants, except CC001). The movements where visually cued with a ∼2 s interval in epochs of 15-30 s, which were alternated with resting epochs. The length of the task in ALS participants ranged between 275 s and 531 s. The length of the task in control participants was 300 s.

### Analysis of structural MRI data

Structural MRI images of all control and ALS participants were processed using the same surface reconstruction pipeline of FreeSurfer 7.1, which includes the creation of surface reconstructions, making local estimates of cortical thickness, and defining regions of interest ^18,19^. The created white matter and pial surfaces were superimposed on the structural image and visually inspected for tissue misclassifications, which were manually corrected if needed. Furthermore, the Desikan-Killiany annotation ^20^ was checked for accurate segmentation of the precentral and postcentral gyrus. The FreeSurfer output was then used to generate Cgrids of the sensorimotor cortex. Cgrids are rectangular and gridded representations of portions of the brain that are created fully automatically by fitting polynomials to the borders between brain areas on a flattened surface, and subsequently interpolating the coefficients of the fit ^21,22^. The sensorimotor Cgrids created in this study covered the precentral and postcentral gyrus as according to the Desikan-Killiany parcellation ^20^ of the FreeSurfer pipeline, with the horizontal axis running from anterior to posterior, and the vertical axis running from ventral to dorsal^40,41^. We visually inspected the Cgrid structure on an inflated surface representation to verify the validity of the procedure. The Cgrid representation was used to facilitate visualization and interindividual comparisons. Grey matter thickness was assessed by averaging the thickness across vertices in Cgrid tiles, while weighing for the surface area of each vertex, and was subsequently averaged over the left and right hemisphere.

A linear regression model was used for fitting the relationship between age and grey matter thickness in the healthy volunteers for each sub-portion of the Cgrid, while also establishing the 95% prediction interval of the normal fit (i.e. the interval enclosing 95% of any new observation).

### Analysis of functional MRI data

The functional volumes were corrected for head motion and coregistered to the surface reconstructed structural images using SPM12 (https://www.fil.ion.ucl.ac.uk/spm/software/spm12/). The alignment between the surface reconstructions and functional images was visually inspected using FreeSurfer’s Freeview. To map functional data to the cortical surface, we used the voxels enclosing the middle layer of the grey matter, and applied surface-based smoothing with a Gaussian kernel of 6 mm FWHM. The resulting Cgrid functional data was then subjected to a first-level analysis in SPM12, generating maps with beta coefficients representing brain activity during (attempted) hand movements for each participant.

We assessed the presence of reductions of the amplitude of the fMRI activity in the motor hand area in ALS participants using the fMRI data acquired during the (attempted) right-hand movement task. We defined the motor hand area based on the group-mean activity of real right-hand movements of control participants using the lowest threshold producing a single cluster in the precentral gyrus. The postcentral gyrus was excluded from these subsequent analyses since control participants had movement-related feedback while the paralyzed ALS participants did not. To avoid that the motor hand area for control participants was established using the same data as for which the amplitude of fMRI activity was estimated (double dipping), the motor hand area was redefined for every control participant using a leave-one-out procedure. This was done by calculating the group-mean activity leaving out the control participant for which the amplitude of fMRI activity was estimated.

The timeseries data obtained from the motor hand area was then high pass filtered with a cutoff at 0.01 Hz to remove noise, and the mean signal across the hand area was calculated and scaled to represent percent signal change. The resulting timeseries was subjected to a multiple regression analysis including a factor for (attempted) hand movements vs rest, yielding a single regressor coefficient reflecting the amplitude of the fMRI activity in the motor hand area and its 95% confidence interval for each participant.

Similarly to the analysis of the grey matter thickness, the effects of age on fMRI activity in control participants were modelled with a linear regression, but now using the normalized inverse of the confidence intervals of the beta coefficients as weights, to account for variations in the standard-deviations of the individual measurements.

### Acquisition of ECoG data

Two distinct iBCI setups were employed: the CortiCom system and the UNP system. Implanted electrodes were always positioned on the left hemisphere.

The CortiCom system ^23^ consisted of subdural ECoG grids with a total of 128 channels that were connected to a percutaneous Neuroport pedestal (Blackrock Neurotech, Salt Lake City, UT, USA). Reference wires were positioned between grids and dura. During neural signal recordings, the pedestal was connected to an external device for signal amplification, digitization, and digital transmission (NSP, Blackrock Neurotech). The grids were surgically placed via a craniotomy over the sensorimotor cortex (Figure 1) based on anatomical landmarks, superficial blood vessels and hotspots of fMRI activity. Participant CC001 received two 64-channel grids (PMT Corporation, Chanhassen, MN, USA) of 8 x 8 electrodes with an inter-electrode distance of 4 mm (exposed electrode diameter 2 mm). Participant CC002 received four 32-channel grids of 4 x 8 electrodes (CorTec Neuro, Freiburg, Germany): the most dorsal and ventral grid had a spacing of 4 mm, while the two middle grids had an inter-electrode distance of 3 mm (exposed electrode diameter 1 mm) (Figure 1). The CortiCom implant was aimed at high-dimensional decoding of speech and hand movements.

**Figure 1:**
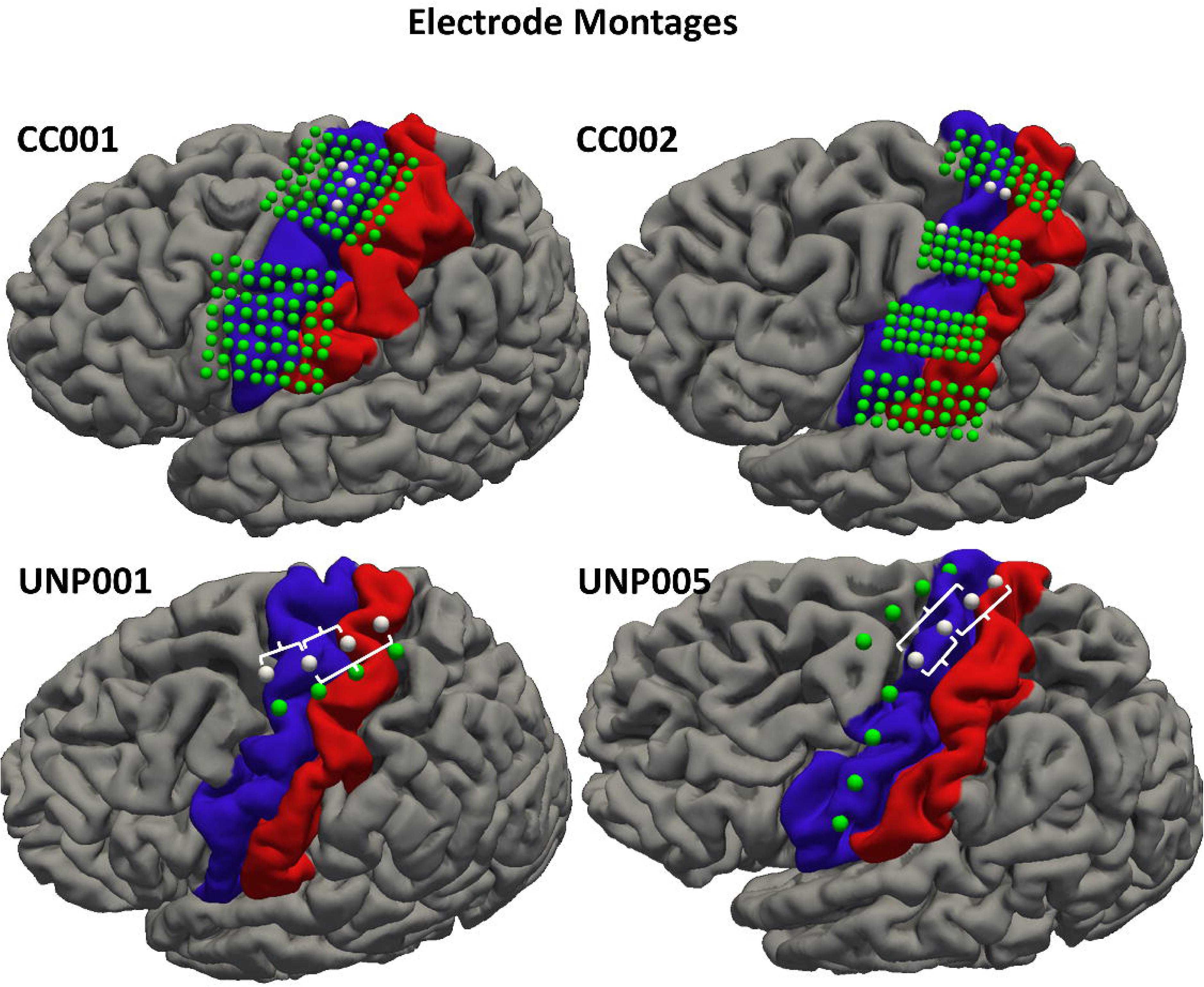
Electrode locations over the sensorimotor cortex. The locations of electrodes of the implanted grids over the sensorimotor cortex superimposed on the pial surface of the left hemisphere of the 4 implanted ALS participants. Electrodes used for the comparison with (f)MRI metrics are colored white, which for UNP001 and UNP005 are also the electrodes connected to the amplifier during the second surgery. Brackets indicate the selected electrode pairs for UNP participants. The prefrontal electrode strips of UNP001 and UNP005 are not shown. The precentral gyrus is represented in blue, and the postcentral gyrus in red.

The UNP system ^16^ consisted of two subdural ECoG strips connected to a subcutaneously implanted amplifier/transmitter device. Each strip contained 4 platinum electrodes (*Resume-II*® or *Subdural Leads*, Medtronic, off-label use) that were recorded in a bipolar montage. The electrodes had an exposed diameter of 4 mm and an inter-electrode distance of 10 mm. The UNP system was surgically implanted in two stages. During the first surgery, 4 strips were implanted through 10 mm burr holes and placed over sensorimotor cortex based on presurgical fMRI, anatomical landmarks and superficial bloodvessels. Measurements obtained at the intensive care unit in the following days were used to identify the 2 ECoG strips that showed the strongest signal changes during task performance. During the second surgery, these 2 strips were connected to the amplifier/transmitter device (Figure 1), which was placed subcutaneously below the left clavicle. For UNP001, two strips were implanted over sensorimotor cortex (one connected) and two over the dorsolateral prefrontal cortex (one connected). For UNP005, three strips were implanted over the sensorimotor cortex (two connected), and one over the dorsolateral prefrontal cortex (none connected). Here, we only use data from the sensorimotor strips. Electrode recordings were made using the Medtronic Activa PC+S system. The UNP device was aimed to provide basic (1-dimensional) yet reliable communication for individuals in a locked-in state.

For UNP001, CC001, and CC002, the included data was acquired in 3 sessions across 3 separate days in the period 2 to 4 months after implantation. While ECoG data was acquired on many more occasions during the following months/years, the three sessions were chosen to ensure full recovery from surgery and familiarization with the task procedures, while minimizing the extent of disease progression relative to the (f)MRI scanning. For UNP005, the onset of ECoG data acquisition was delayed by 2 months due to COVID lockdown restrictions, and therefore we included data from three sessions acquired between 4 to 6 months after implantation.

UNP001, UNP005, and CC002 performed a task in a blocked design that alternated active epochs with resting epochs. In the active epochs, participants continuously attempted hand movements at their own pace. The length of each epoch was 15s. CC002, UNP001, and UNP005 performed a total of 24, 18, and 60 epochs respectively. CC001 conducted a task in an event-related design in which he was instructed to move his hand once after each 100 ms cue. He then rested for the remainder of the 3-6s uniformly distributed inter-trial interval. He performed 150 trials (with 30s rest after each 50 trials) in the first session and 90 trials (with 30s rest after each 30 trials) in the second and third session. Since he was cued to attempt one movement at the time, we defined an active epoch as 0.3-0.8s after cue onset, and a resting epoch as 0.8-0.3s before cue onset. Synchronization of task and ECoG signals was accomplished through triggers from the task to acquisition PC, or running task and signal recordings on the same computer using software derived from BCI2000 ^24^.

All participants included in the ECoG reference group were implanted with clinical grids (1 cm center-to-center electrode distance, 2.3 mm exposed surface diameter, AdTech, Racine, USA) covering, among other regions, the motor hand region. ECoG signals from the clinical grids were acquired with a 128-channel Micromed recording system (Treviso, Italy; hardware band-pass filter 0.15-134.4 Hz) at 512 Hz. Participants performed a task where epochs (mean/SD epoch duration=23.8/7.3 s) of finger-tapping with the hand contralateral to the implanted grid were alternated with resting epochs of equal length. The total task duration varied across participants and ranged from 4.3-21 minutes (Mean/SD total duration=10.4/7.3 min).

### Analysis of ECoG data

ECoG data was sampled at 200 Hz for the UNP participants, at 1000 Hz for the CortiCom participants, and at 512 Hz for the ECoG reference group. For analysis, the data of the CortiCom participants was down sampled to 250 Hz. Line noise (50 Hz or 60 Hz for measurements done in Europe and the USA, respectively) was removed using a band-stop filter. The data of CortiCom participants and the ECoG reference group was common-average re-referenced, excluding bad channels. Subsequently, a Gabor wavelet decomposition was applied in 1 Hz bins with decreasing window length (4 wavelength full width at half maximum) to establish the changes in the high frequency band (HFB, 65-95 Hz) and the low frequency band (LFB, 8-30 Hz) power over time. The band power was averaged across frequency bins and then across the time samples in each (attempted) movement or resting epoch, resulting in a single value per epoch and electrode (pair). Then the correlation coefficients between the band power per epoch and an on/off vector were calculated, which were subsequently transformed to Fisher’s z-scores to make the correlation results more normally distributed and allow for the calculation of 95% confidence intervals. The Fischer z-scores were taken as metric for ECoG signal quality. We used this correlation-derived metric because it reflects the extent to which task-related band-power modulation can be distinguished from non-task-related variability, which is directly relevant for BCI control.

For participant CC001 the band power was averaged per 6 trial epochs before calculating correlations, to ascertain that the durations of the epochs used for calculating the band power was similar across participants. This avoided bias in the reliability of the power estimates and thus in the amplitude of the Fisher z-scores.

Electrode locations of all implanted individuals (ALS participants and the ECoG reference group) were determined using a head-CT scan acquired after implantation for all subjects except CC002. Using the ALICE pipeline ^25,26^, electrode locations were extracted from the CT scan and projected onto the pial surface reconstruction of each participant, and subsequently to the Cgrid. Electrode locations for CC002 were determined using the Gridloc toolbox ^27^ due to a brain shift after implantation.

For CortiCom participants, we selected the 3 electrodes with the highest z-score for HFB fluctuations during (attempted) movements vs rest that were also over the focus of highest fMRI activity during the movement vs rest contrast in the imaging control group. For UNP participants, where data acquisition involved pairs rather than single electrodes, 3 electrode pairs with the highest z-score for HFB fluctuations for (attempted) movements vs rest were selected, with the prerequisite that one of the electrodes of each pair was located over the focus of highest fMRI activity during the movement vs rest contrast in control participants. The electrode-selection procedure for the ECoG reference group was similar to that of CortiCom participants, with the exception that we only selected the single electrode with the maximum HFB fluctuations in the fMRI activity hotspot of control participants. Only a single electrode was included because the absence of navigation of grid placement based on fMRI in combination with the large interelectrode distance, made multi-electrode coverage of the motor-hand area unlikely. The Z-s The effects of age on ECoG activity was modelled with a linear regression.

### Second level analysis

For establishing the relationship between ECoG on the one hand, and fMRI and grey matter thickness on the other, we first standardized the measured ECoG responses, grey matter thickness, and fMRI activity for each ALS participant to Z-values, based on the distribution of control subjects using the formula:

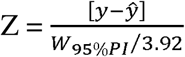

Where y is the measured value, the predicted value based on the linear age model, and W_95%PI_ the total width of the prediction interval at the age of the participant. For calculating the Z-values for grey matter thickness in CC001 and CC002 we used 0.14 as the denominator in the formula, instead of the observed width of the predication interval in the participant’s age. The value of 0.14 is an estimate of the (age-corrected) standard deviation across individuals for grey matter thickness in the precentral gyrus based on multiplatform consortium study ^28^. The reason for this alternative benchmarking for CC001 and CC002 was that their MRI data was acquired on different scanner platforms at the two sites, and platform effects could bias towards a larger magnitude of the Z-values when normalizing using data that was acquired in a single platform. After the calculation of the Z-values, we calculated the Z-value for the difference (Δ*Z*) for each metric between the two ALS participants within each system (CortiCom/UNP):

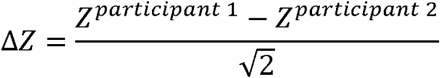

We defined the second-level test-statistic as the inner product of ECoG on the one hand and grey matter thickness/fMRI activity on the other:

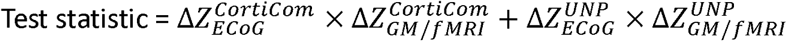

To estimate the likelihood of by chance (one-sided) obtaining a test-statistic equal or larger than the observed test-statistic, we performed a Monte-Carlo simulation (one million repetitions) where the different ΔZ values were randomly selected from a normal distribution with a mean of 0 and standard deviation of 1.

## Results

### Structural MRI results

We assessed grey matter thickness in the precentral and postcentral gyri of the sensorimotor cortex in six individuals with advanced ALS and compared their data to the healthy controls. In healthy controls, cortical thickness in the precentral gyrus showed a clear linear decline with age. Four of the six ALS participants (CC002, CC003, UNP005, and UNP006) demonstrated significant cortical thinning in the precentral gyrus, which was defined as falling outside the 95% prediction interval of age-matched controls, while the postcentral gyrus remained within the normal range for all ALS participants (Figure 2). The left and right precentral gyri were similarly affected in individuals with ALS (r=0.978; p<0.001), suggesting bilaterally symmetric disease involvement, justifying the averaging across hemispheres.

**Figure 2:**
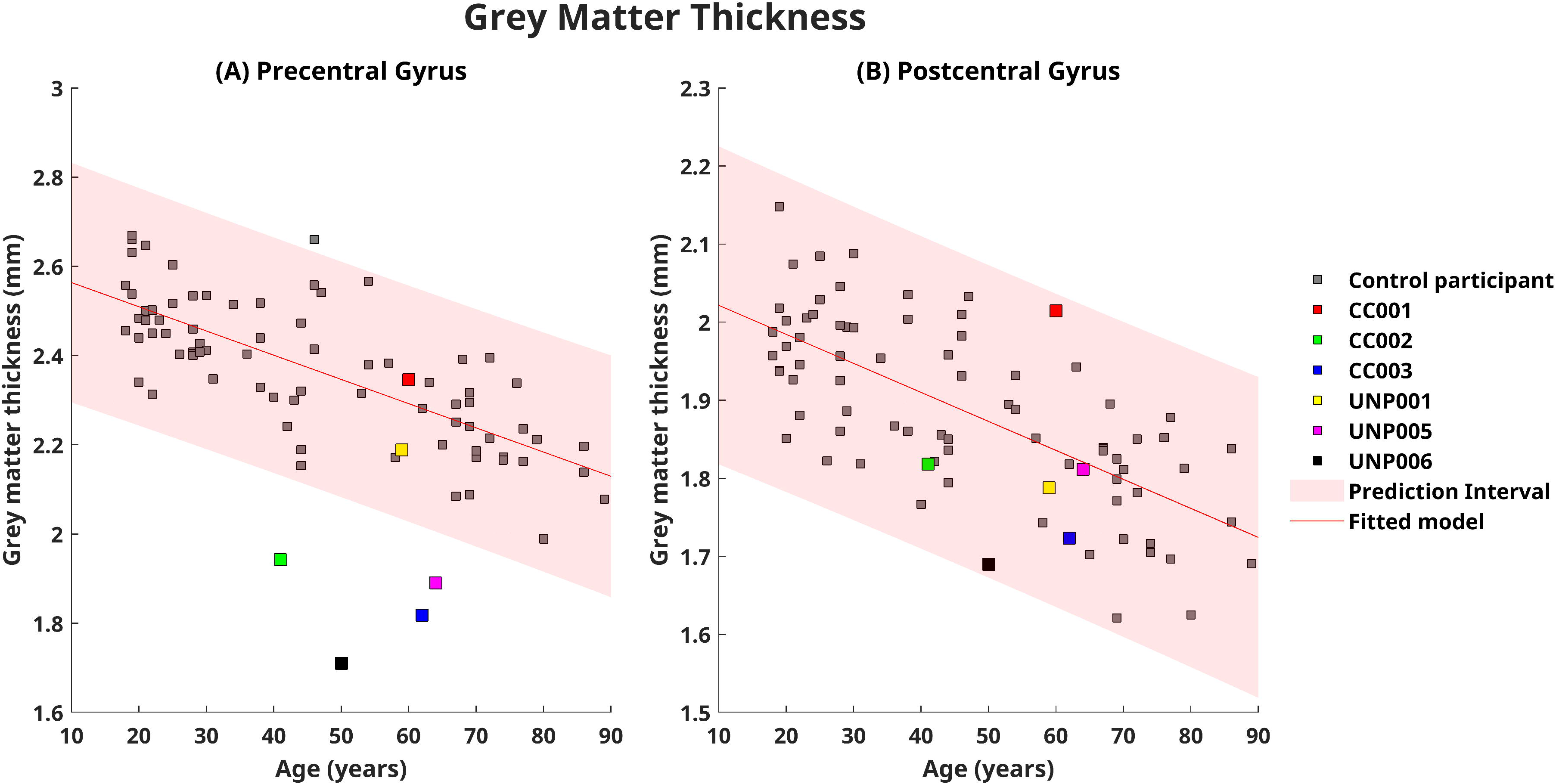
Grey matter thickness in the sensorimotor cortex. Scatterplots comparing grey matter thickness between ALS participants and controls across different ages for the mean of the left and right (A) precentral gyrus and (B) postcentral gyrus. The linear model based on the data of the controls is depicted by a straight red line, which is surrounded by a shaded red area representing the 95% prediction interval. All ALS participants outside the prediction interval significantly deviated from controls. The source data for Figure 2 is in Supplementary Data 2.

We next mapped the cortical thickness to the Cgrid representation of the sensorimotor cortex ^29,30^, allowing for more detailed visualization of regional thinning. In the four ALS participants with significant precentral thinning, reductions spanned nearly the entire precentral gyrus, but largely spared the most ventral portion (Figure 3).

**Figure 3:**
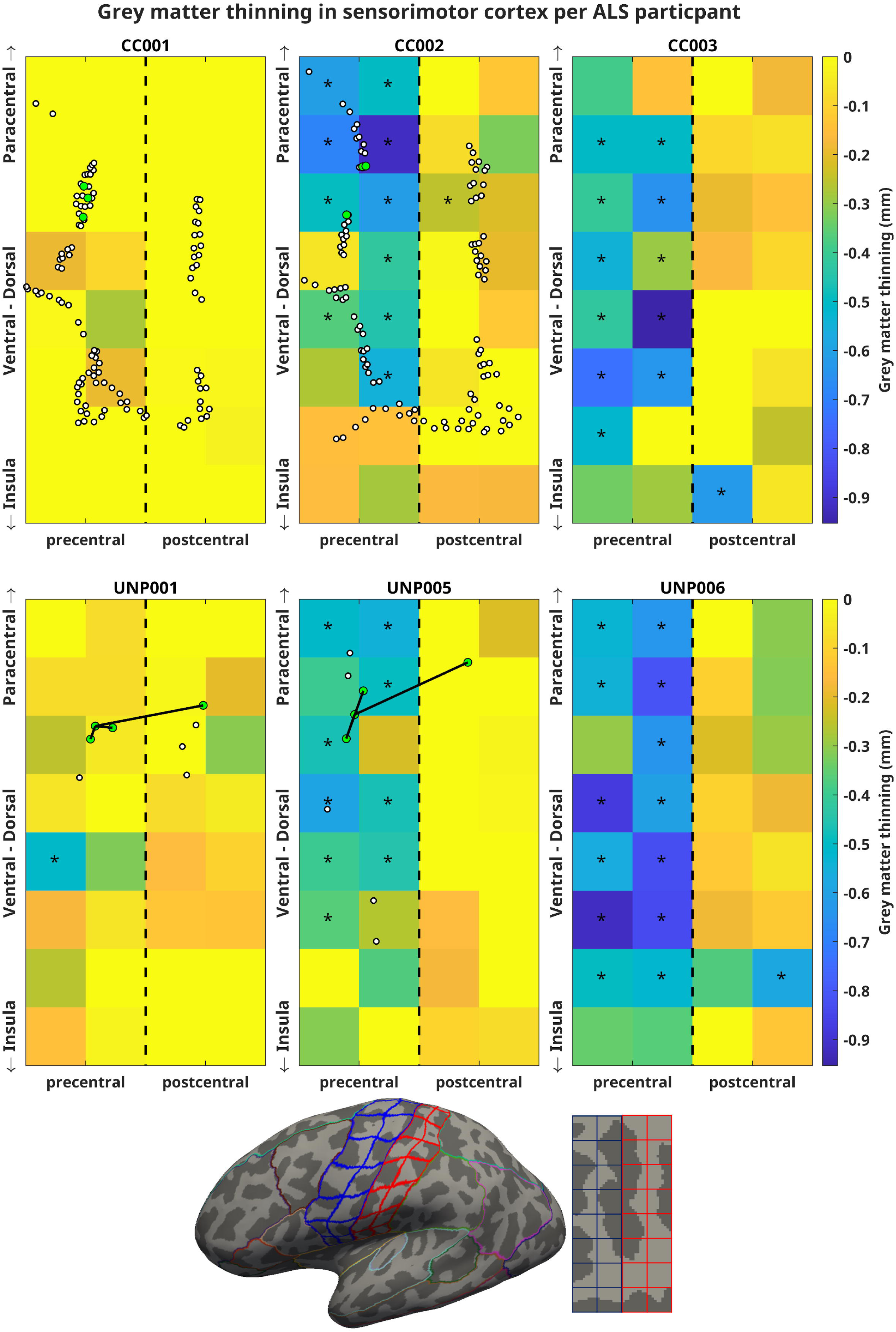
Regional grey matter thinning in ALS participants. Grey matter thinning in the mean of the left and right sensorimotor cortex relative to the model for control participants per tile of a 4×8 Cgrid for the 6 ALS participants. The color index is capped at 0 mm tissue loss for optimal visualization. Asterisks depict reductions outside the 95% prediction interval of controls (uncorrected for multiple comparisons). Green circles depict the electrodes that were used for the comparison with ECoG data, with for UNP001 and UNP005 black straight lines connecting the three selected electrode pairs. The small white circles depict the locations of remaining electrodes. The black dashed vertical straight line in the middle of each grid aligns with the bottom of the central sulcus, i.e. the border between the precentral and postcentral gyrus. The Cgrids are bordered by the insula and paracentral lobule at the ventral and dorsal side, respectively. The source data for Figure 3 is in Supplementary Data 3.

### Functional MRI results

Functional MRI was during either executed or attempted right-hand movements in the left motor hand area. The ALS participants showed marked variability in task-related activation (Figure 4). Significant (p<0.05, FWE-corrected) fMRI responses in the precentral motor hand area were observed in four ALS participants (CC001, CC003, UNP001, and UNP005), while three (CC002, CC003, and UNP006) exhibited amplitudes below the 95% prediction interval of controls when accounting for age (Figure 5).

**Figure 4:**
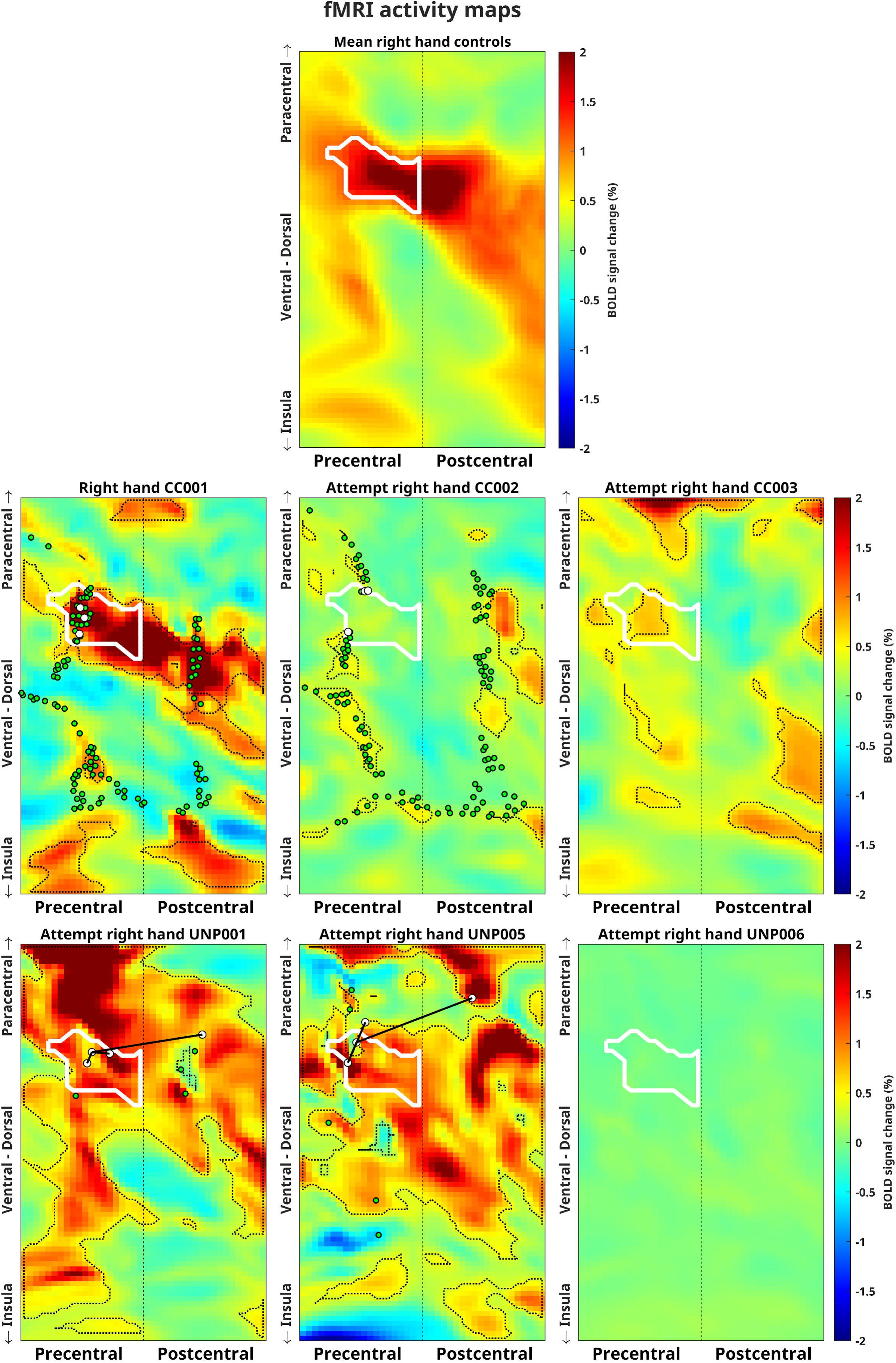
fMRI activity in the sensorimotor cortex. fMRI activity maps in Cgrid space of the left sensorimotor cortex during executed right-hand movements (group-mean healthy controls) and (attempted) right hand movements (6 ALS participants). Activity is expressed in percent signal change. Areas in ALS participants with significant activity (p<0.05; FWE corrected) are encircled by a dotted black line. The white line encircles the precentral motor hand area of interest, as defined on the group-mean activity of the healthy control participants. The vertical dotted straight line in the middle of each half aligns with the bottom of the central sulcus, i.e. the border between the precentral and postcentral gyrus. The Cgrids are bordered by the insula and paracentral lobule at the ventral and dorsal side respectively. White circles depict the electrodes that were used for the comparison with ECoG data, with for UNP001 and UNP005 black lines connecting the three selected electrode pairs. Green depicts the electrodes excluded from subsequent analyses. The source data for Figure 4 is in Supplementary Data 4.

**Figure 5:**
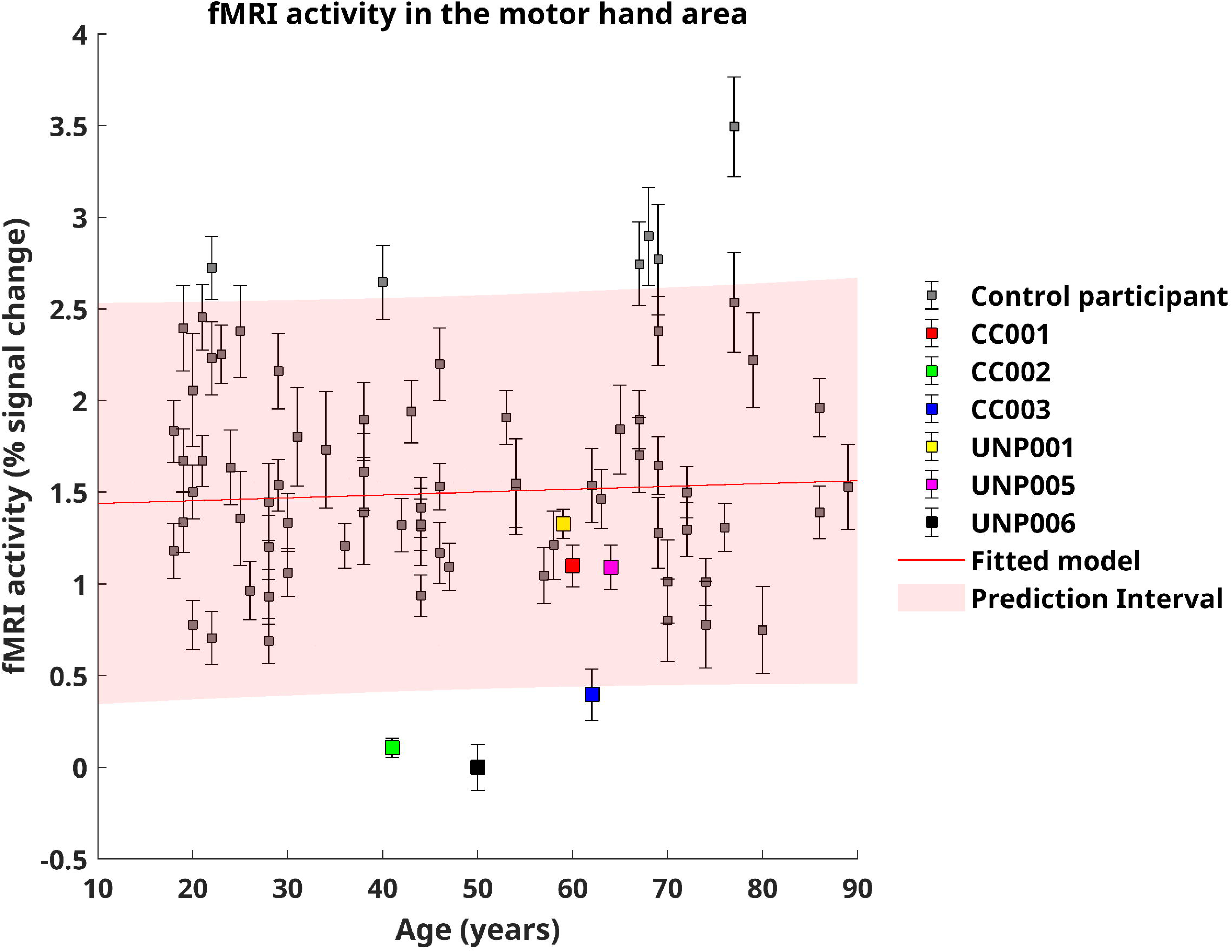
fMRI activity in the motor hand area. Scatterplot of age vs fMRI activity in the motor hand area in controls during real movements, and in ALS participants during (attempted) movements. The first-order polynomial fit on the data of the healthy controls is depicted by the straight red line, which is surrounded by a shaded red area representing the 95% prediction interval. Error bars reflect the 95% confidence intervals of the activity estimates. The source data for Figure 5 is in Supplementary Data 5.

### ECoG results

We analyzed electrocortigraphy (ECoG) recordings from the four ALS participants who were implanted with either grid-based (CortiCom; CC001, CC002) or strip-based (UNP; UNP001, UNP005) systems over the left sensorimotor cortex, and from our control group of 24 individuals with epilepsy. ECoG recordings were made during (attempted) hand movements tasks.

The HFB and LFB responses of the selected electrodes over the motor hand area in ALS participants are shown in Figure 6. The mean ECoG quality across selected electrodes and electrode-pairs of ALS participants was compared to that of controls (Figure 7). CC001 and UNP001 had above average, and CC002 and UNP005 below average HFB ECoG signal quality (Figure 7), albeit not significantly different from controls on a single subject basis. LFB ECoG quality only marginally deviated from that of controls.

**Figure 6:**
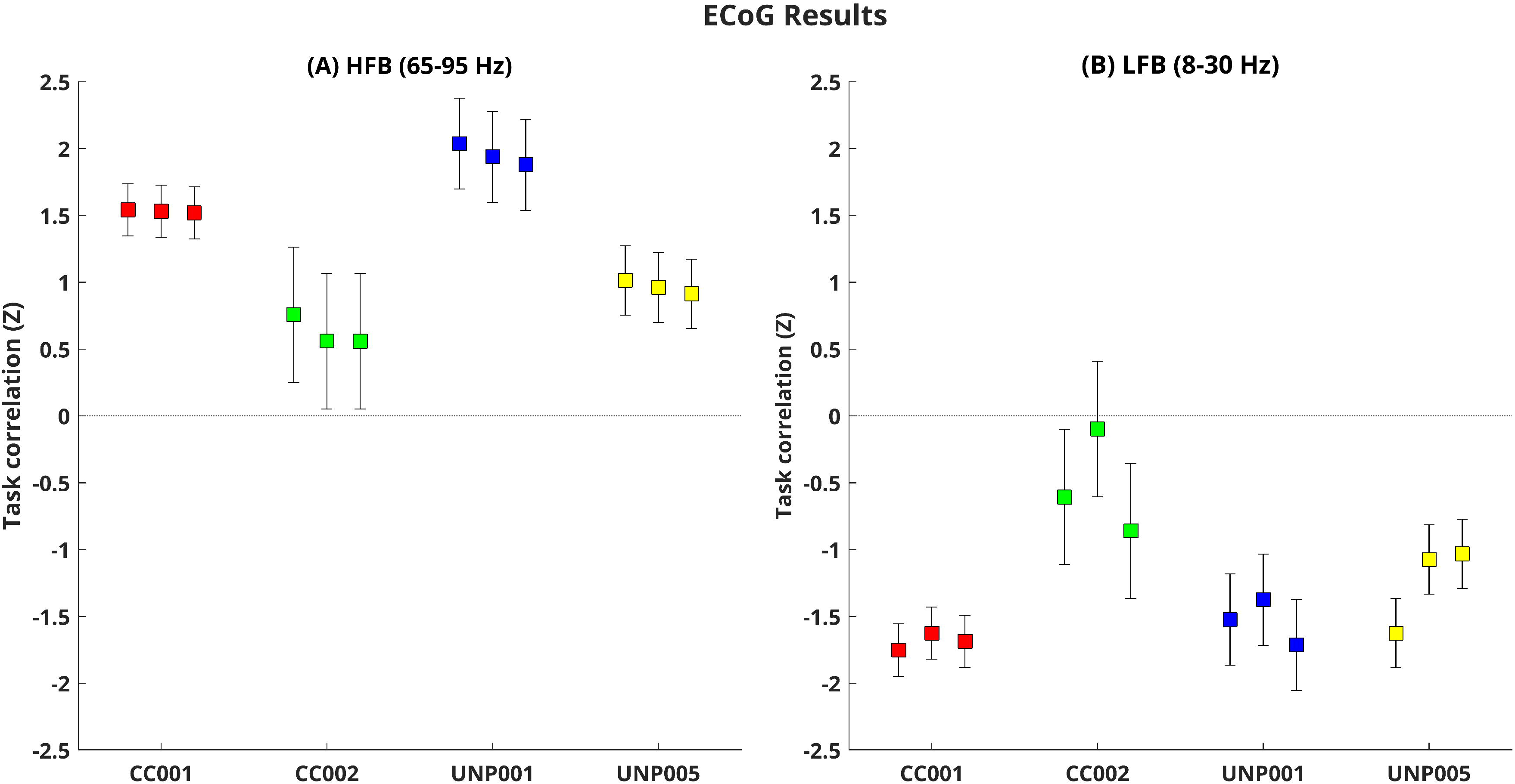
ECoG signal quality during attempted movement. Results of the ECoG measurements, showing the Fisher’s z-scores of the of the selected 3 electrodes (CC001 & CC002) or 3 electrode pairs (UNP001 & UNP005) with the highest task correlation for the HFB power in the vicinity of the fMRI hotspot of controls in the precentral gyrus. Error bars indicate the first-level 95% confidence intervals. Panel (A) shows the z-scores for the HFB power (65-95 Hz), and panel (B) for the LFB power (8-30 Hz). For the LFB more negative responses reflect a stronger response.

**Figure 7:**
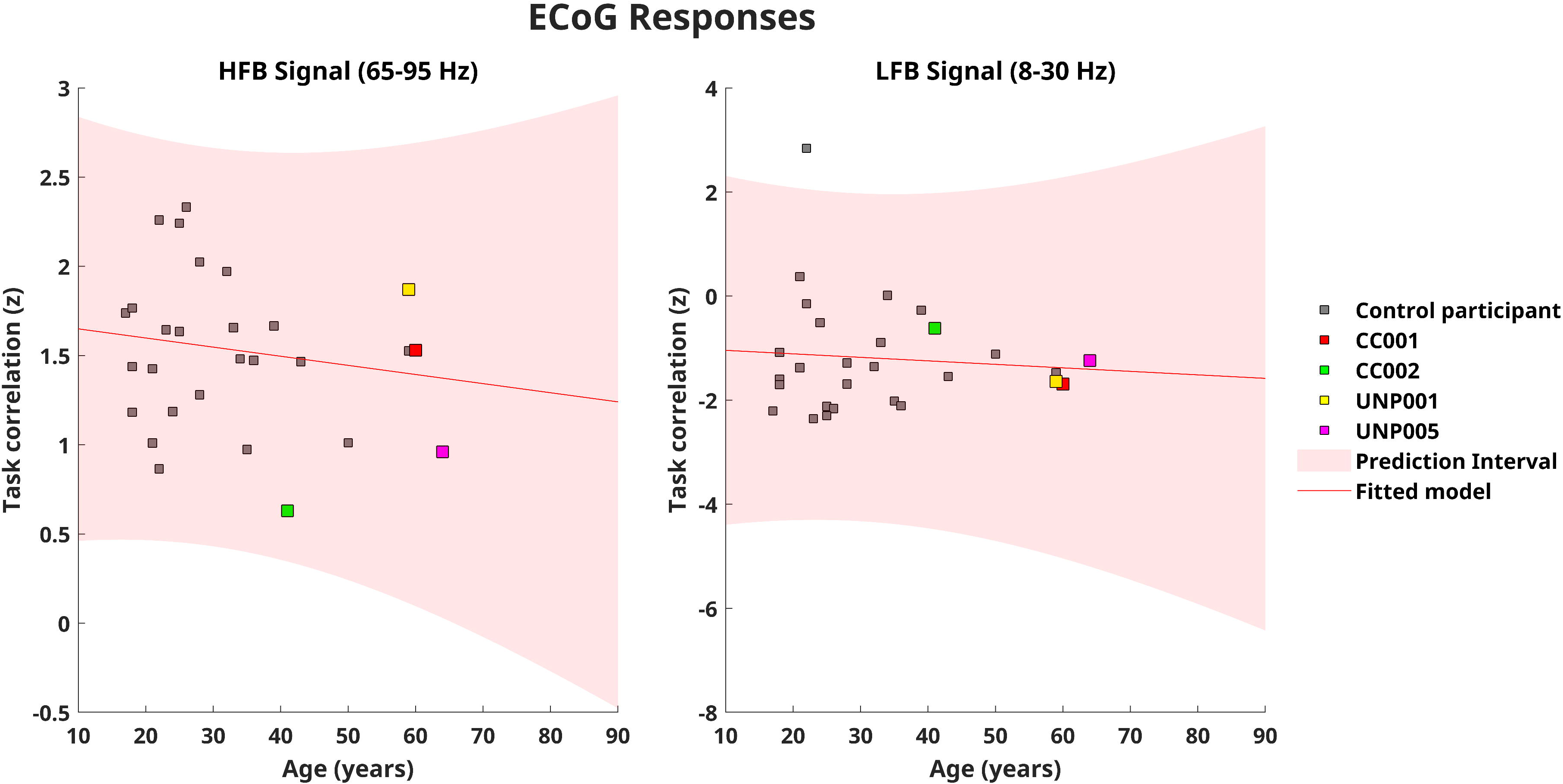
ECoG signal quality relative to age. Scatterplots comparing ECoG quality between ALS participants and controls across different ages for (A) the HFB (B) the LFB. The linear model based on the data of the controls is depicted by a straight red line, which is surrounded by a shaded red area representing the 95% prediction interval. The source data for Figure 7 is in Supplementary Data 6.

Power spectral density during active and resting periods is shown for the implanted ALS participants and for the mean of the ECoG reference group in figure 8. Visual inspection revealed that while three of the implanted ALS participants showed responses that were qualitatively similar to that of the control group, one participant (CC002) lacked both the HFB and LFB differences associated with active and rest.

**Figure 8:**
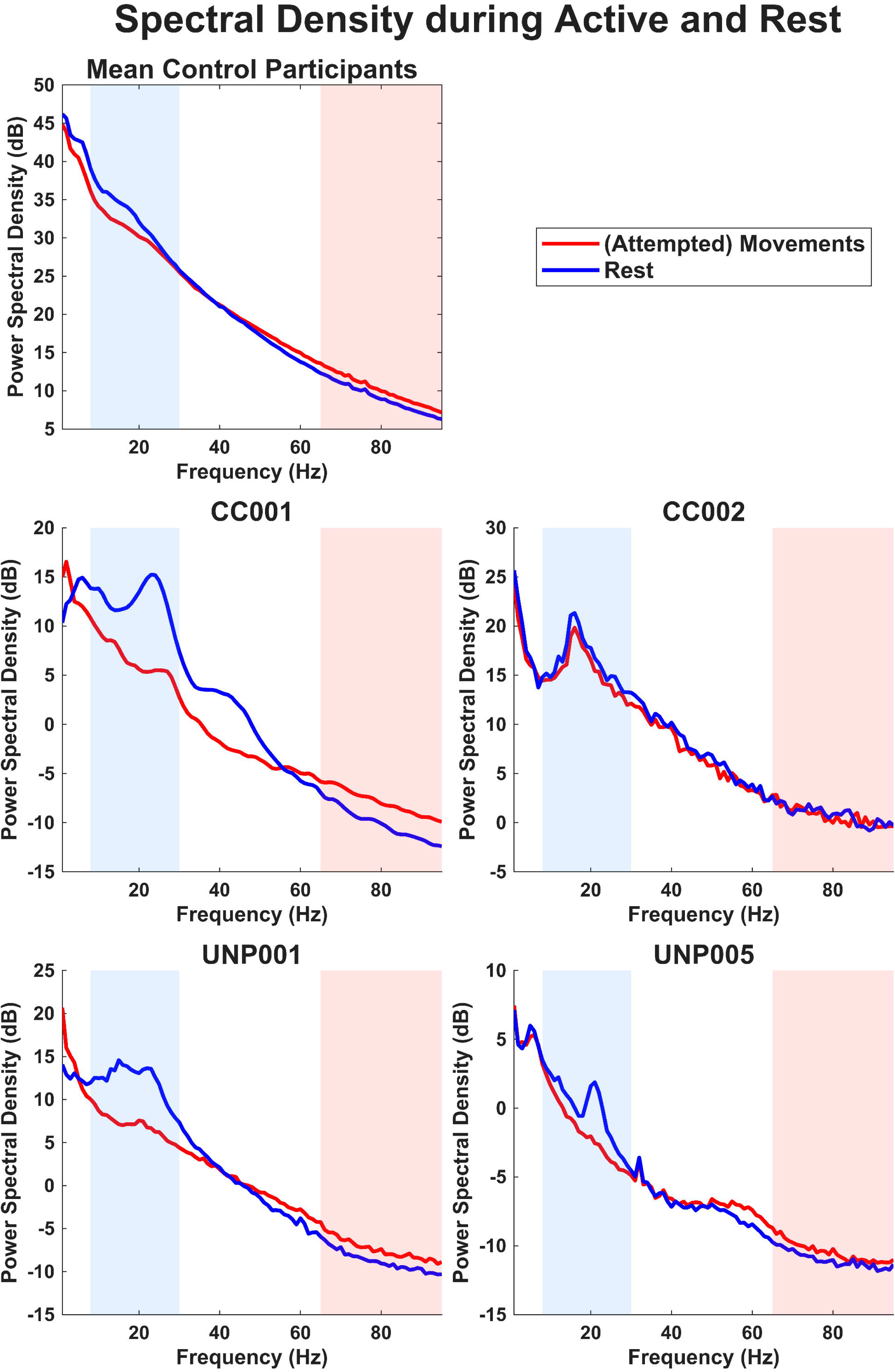
Spectral Densities for ALS participants and Controls. Differences in the spectral density between active and rest periods for the control participants and for every implanted ALS participant. The blue and red shaded area represents the LFB and HFB respectively. The source data for Figure 8 is in Supplementary Data 7.

### Second level analysis

To test the relationship between ECoG quality on the one hand, and grey matter thickness and fMRI activity on the other, we standardized the deviations of ALS participants from the control populations to Z-values for all metrics (Table 1). Then we calculated the Z-values for the difference (ΔZ) for each metric between the 2 implanted ALS participants that were measured with the same system (CortiCom/UNP) (Table 1). Only pairwise comparisons were made to negate potential bias by the ECoG implant system. Using a Monte Carlo simulation, we tested the likelihood that the observed alignment of ΔZ values across different metrics could have occurred by chance. This test revealed a significant association of HFB ECoG signal quality with grey matter thickness (p=0.01), but not with fMRI activity (p=0.10). LFB ECoG signal quality was not associated with grey matter thickness (p=0.83) or fMRI activity (p=0.70).

### Comparing fMRI activity and grey matter thickness

The fact that reduced signal quality was associated with grey matter thinning, but did not align with reduced fMRI activity is notable. To investigate whether this discrepancy could be attributed to differences in the sensitivity of the two metrics (cortical thickness and fMRI activity) for detecting abnormalities, we compared their detection thresholds. Visual inspection of the amplitude of the fMRI activity in healthy controls revealed substantial variation, with a 95% prediction interval ranging from 0.3% to 2.6% signal change (Figure 4). Assuming that disease processes lead to proportional reductions in fMRI activity relative to a healthy state, we simulated detection rates by incrementally decreasing amplitude of fMRI activity in controls from 0% to 100%, in 1% steps. At each step, we calculated the proportion of measurements falling outside the 95% prediction interval that is depicted in Figure 4. Only reductions of 87% or more had a detection rate above 95%, indicating that reductions in fMRI activity can only be reliably identified in cases of near-complete breakdown of the task response. In contrast, grey matter thickness abnormalities were detected in 95% of cases when reductions exceeded 19%. This stark difference in sensitivity between the two metrics suggests that the absence of detectable reductions in fMRI activity relative to controls does not mean absence of reductions relative to his previous healthy state (see Figure 1 in Supplementary Data 1) for detection rates for all percentual reduction steps).

## Discussion

This study aimed to investigate the relationship between neural signal features used for ECoG-iBCI performance in people with ALS based and presurgical imaging results. Four ALS participants underwent BCI implantation. There was significant thinning of the grey matter in the precentral gyrus in two of them. This grey-matter thinning was associated with reduced HFB but not LFB ECoG responses during (attempted) movements. The relationship between the presurgical fMRI results and signal quality was less clear. The current results provide preliminary evidence that grey matter thinning in the precentral gyrus may serve as a useful imaging-based predictor of eligibility for motor-based iBCI.

Grey matter thinning in the motor cortex of individuals with ALS is well documented ^31–33^. Furthermore, a correlation between disease progression and grey matter thinning in the motor cortex has been reported, suggesting that more severe atrophy is likely as ALS advances ^34^. Indeed, the degree of thinning in four of our late-stage ALS participants (approximately 0.4 mm compared to age-matched controls) exceeded the most severe thinning reported in a cohort of 112 earlier-stage individuals with ALS (approximately 0.3 mm) ^31^. It is thus likely that grey matter tissue loss is more severe for individuals in more advanced disease stages and in those who approach a locked-in state.

Grey matter loss in the primary motor area (M1) of participants with ALS is thought to primarily reflect effects on the motor neurons in layer V, an output layer with projections to the brainstem and spinal cord that are crucial for initiating voluntary motor movements ^35^. Degeneration in this layer may significantly impact ECoG-based iBCIs, as an animal model indicates that approximately 85% of the HFB power fluctuations detected by ECoG originate from the pyramidal cells in layer V and VI ^36^. This could explain the relatively low signal quality observed in the two ALS participants with marked cortical thinning in M1, but it remains uncertain whether layer V is solely affected.

Our findings provide preliminary evidence that people with ALS can benefit from an ECoG-based iBCI, as long as the grey matter in the precentral gyrus retains sufficient integrity to generate motor-related activity. ALS is a highly heterogeneous disease, with varying degrees of upper and lower motor neuron involvement across individuals and these patterns can change over time ^37^. Consequently, the success of iBCI functionality will likely depend on individual characteristics, including the cortical thickness at the time of implantation. In the present cohort, two of six participants showed preserved precentral grey matter integrity together with successful iBCI use, corresponding to 33.3%, but with a wide Wilson 95% confidence interval of 9.7–70.0%. This uncertainty, together with the highly selected nature of the cohort, precludes a population-level estimate of the fraction of individuals with ALS who may benefit from an iBCI.

In addition, we expect grey matter thinning to impact iBCI performance over time, even in individuals with ALS whose precentral gyrus appears unaffected at the time of implantation. Longitudinal MRI studies consistently show that cortical thinning in the precentral gyrus is a progressive feature of ALS and correlates with upper motor neuron burden and clinical decline ^38–40^. In addition, in ALS participant UNP001, iBCI performance, while initially high, declined in the course of seven years after implantation, with a CT scan revealing significant brain tissue loss over time ^41^. By contrast, participant CC001, whose precentral grey matter thickness was preserved relative to age-matched controls, had been using the implant for approximately two years at the time of writing. Thus, although our results show that individuals with ALS can benefit from an iBCI for a substantial time period, a decline of the source signals from the precentral grey-matter may eventually reduce iBCI performance^31^. These observations support the need for predictors, e.g. clinical and genotypical characteristics, for precentral grey-matter integrity in late disease stages. Not only could these help to assess eligibility for an iBCI, but they could also facilitate the assessment of the size of the target population for this technology.

The relationship between fMRI activity levels in the motor hand area and ECoG signal quality seems more complicated. A closer examination revealed considerable between-participant variation in the amplitude of fMRI activity in healthy controls, with the 95% confidence interval ranging from approximately 0.3% to 2.6% signal change. This finding is consistent with previous research showing substantial inter-individual differences in the amplitude of fMRI activity ^42^. Based on this variability, we estimate that an average reduction in the amplitude of fMRI activity of at least 87% relative to the healthy state is required to be detected with 95% confidence. This suggests that fMRI can most likely successfully predict cases of reduced ECoG signal quality, but only if these coincide with a vast attenuation of fMRI activity. This contrasts with grey matter thickness measurements, which can reliably detect changes as small as 19%. Note, however, that fMRI can still play a pivotal role in determining the optimal location of the implant ^43^, as, although the amplitude of fMRI activity can vary substantially, the spatial patterns thereof within individuals are highly consistent ^44^.

A key limitation in the interpretation of the current results is the small sample of ALS participants with iBCIs. Our second level test derives inference by benchmarking ALS measurements to control derived normative distributions. This assumes that, after removing covariance between ECoG signal quality and imaging metrics, the standardized deviations (Z scores) in ALS are commensurate with those of controls. If dispersion in ALS is larger or tails are heavier the significance of our findings may be inflated. We therefore interpret the observed association between precentral thickness and HFB ECoG as provisional and conditional on this assumption. However, violation of this assumption would imply that some individuals with ALS have significantly reduced grey matter thickness without this affecting ECoG quality. We consider this unlikely, as the detected grey matter abnormalities are observed in the area from where we acquired the ECoG signals ^17^. Additional but more indirect evidence for a relationship between grey matter thickness and ECoG quality comes from the two non-implanted ALS participants. These two participants exhibited significant reductions in both grey matter thickness and fMRI activity. Given the established relationship between HFB ECoG responses and fMRI activity ^45^, it is unlikely these participants would have shown unaffected ECoG signals if they had been implanted. Furthermore, various other factors varied across ALS participants (device characteristics, tasks) but by standardizing the measures for comparisons as much as possible, we believe we minimized the effects.

A further limitation concerns the spatial sampling of the ECoG reference dataset. These data were acquired from clinical epilepsy grids that were implanted for clinical purposes rather than to systematically cover the motor hand area. As a result, the selected electrode in each reference participant may have varied in its proximity to the individual motor hotspot. Given the 1 cm inter-electrode spacing of these grids, this sparse sampling likely increases between-participant variance in HFB and LFB signal quality and may reduce sensitivity to age-related effects in the reference group. Importantly, this added variance would be expected to make the normative benchmark more conservative for detecting abnormalities in ALS participants, rather than producing spurious group deviations.

This report is limited to ECoG signals but may have implications for electrical potential-based iBCIs in general, in particular those utilizing signals from intracortical electrodes (e.g. Blackrock Neuroport arrays), endovascular electrodes ^46^ or from scalp-EEG ^47^. All these techniques have extracted meaningful information from sensorimotor signals that may exhibit comparable effects of disease progression as observed in this report. The results we report here are likely to extend to ECoG electrodes in general since they typically record from 10^4-10^6 neurons contributing to the net electrical potential ^48^. Micro contacts such as MEA’s and shanks record from one to a few neurons. Presumably the strongest signals in motor cortex are provided by pyramidal cells ^49^, hence we would expect degeneration of pyramidal cells in ALS to affect BCI performance across the different electrode types. Indeed, decoding performance with Utah arrays in late-stage ALS participants appears to be affected as well (personal communication)

In conclusion, this study highlights the potential of grey matter thinning in the precentral gyrus to serve as a predictor of iBCI performance in individuals with ALS. Based on our data, we conclude that the predictive power of fMRI is limited to cases where activity in the motor cortex is nearly absent. These findings do however need to be confirmed using larger samples to allow for statistical tests that require fewer assumptions. Upon confirmation in a larger cohort, these findings can be used for stratification of patients during inclusion of clinical trials and help to restrict implantation of iBCIs to individuals with ALS, or to disease stages, where they are most likely to offer a substantial improvement in communication ability and quality of life.

## Data availability

The data that support the findings of this study are available from the corresponding author upon reasonable request.

## Data Availability

All data produced in the present study are available upon reasonable request to the authors

## Acknowledgements

We express our deepest gratitude for the ALS participants and their families for their participation.

