## Supplementary data 1 for "Association between motor cortex grey matter loss and inability to control an ECoG-based implanted Brain-Computer Interface in ALS"

| **Patient** | CC001 | CC002 | CC003 | UNP001 | UNP005 | UNP006 |
| --- | --- | --- | --- | --- | --- | --- |
| **Sex** | M | F | F | F | M | M |
| **Informed consent** | Jul. 2020 | Dec. 2023 | Oct. 2024 | Sep. 2015 | Sep. 2019 | Sep. 2020 |
| **Age at inclusion** | 60-64 | 40-44 | 60-64 | 55-59 | 60-64 | 50-54 |
| **ALS type** | Bulbar | Spinal | Spinal | Spinal | Spinal | Spinal |
| **ALSFRS score at inclusion** | 26 | 1 | 1 | 2 | 2 | 0 |
| **Residual muscle control at inclusion** | Several,  limited hand control | Eye movements | Eye movements &  blinks Facial expressions | Eye movements &  blinks Facial expressions | Eye movements &  blinks Facial expressions | Right corner  of the mouth |
| **Communication** | Various; see^1^ | Eye tracker Eye movements  for y/n | Eye movements for y/n | Eye tracker | Eye tracker Mouth movements for y/n | Mouth movements for y/n |

Supplementary table 1: Characteristics of included participants with ALS

|  | **Functional** | | | | **Structural** | |
| --- | --- | --- | --- | --- | --- | --- |
| **Participants** | All UNP | CC001 | CC002&CC003 | Controls | CC001 | All other |
| **Scan type** | PRESTO | EPI | EPI | EPI | MRAGE | MPRAGE |
| **Voxel size (mm)** | 4.0x4.0x4.0 | 3.4x3.4x3.3 | 2.3x2.3x2.5 | 2.9x2.9x3.0 | 1.0x1.0x1.0 | 1.0x1.0x1.0 |
| **Scantime per volume (s)** | 0.608 | 0.8 | 1 | 2.5 | 287 | 356 |
| **TR/TE (ms)** | 23/31 | 800/35 | 1000/25 | 2500/39 | 1640/8.4 | 2500/2.9 |
| **FA (degrees)** | 10 | 52 | 65 | 75 | 8 | 8 |
| **Scanner** | Philips | Siemens | Philips | Philips | Siemens | Philips |

Supplementary table 2: Parameters of the used pulse sequences

1.	Luo, S. *et al.* Stable Decoding from a Speech BCI Enables Control for an Individual with ALS without Recalibration for 3 Months. *Advanced Science* 10, 2304853 (2023).

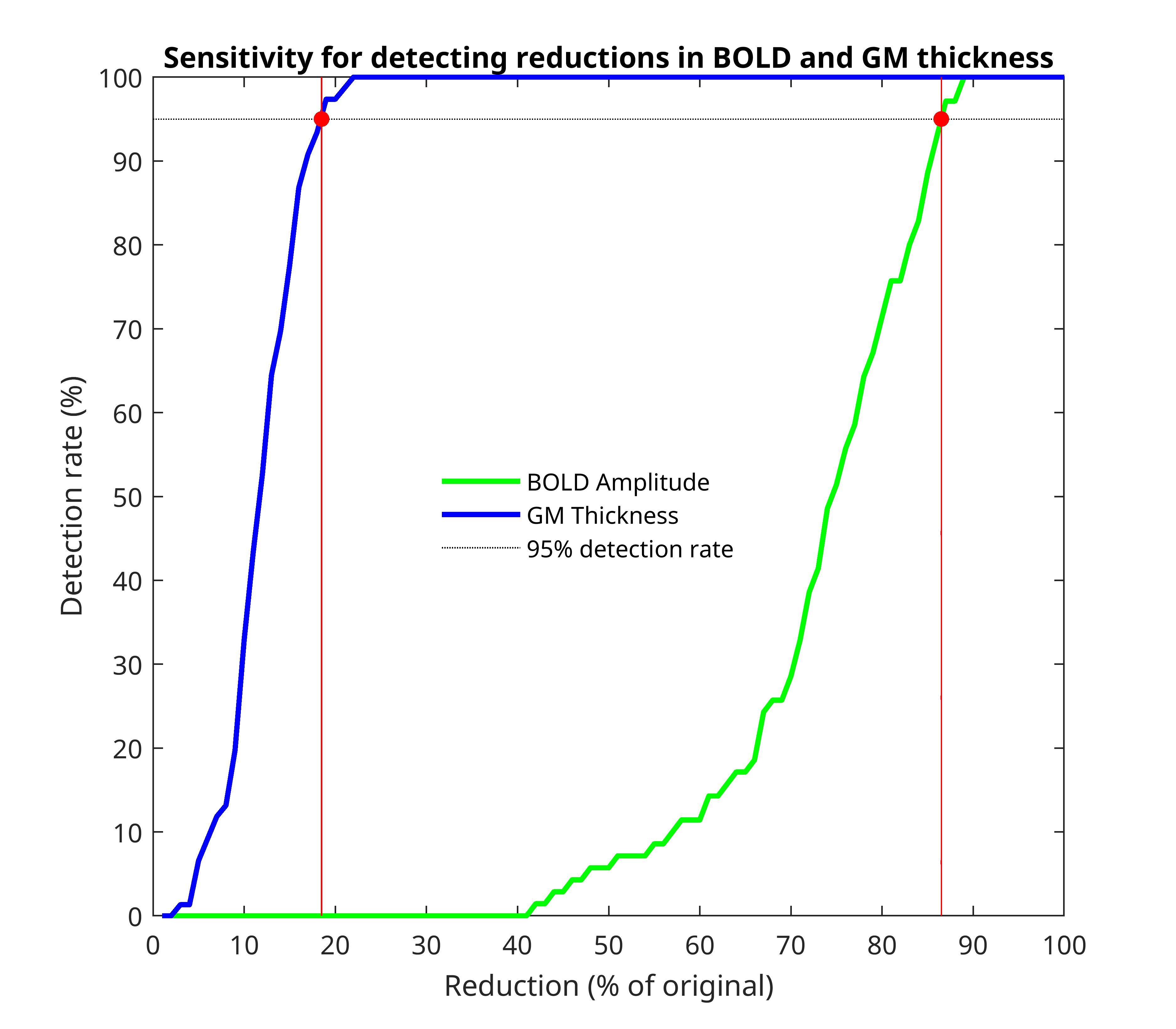

Supplementary Figure 1: Plot depicting the sensitivity for detecting reductions in BOLD amplitudes (green line) and grey matter thickness (blue line). Reductions are expressed as a percentage of the original response. The vertical red lines indicate the reductions at which the detecting chances exceed 95%.
